# Motivators and Barriers to PA Preceptorship in North Carolina

**DOI:** 10.64898/2026.02.16.26346405

**Authors:** Kimberly Stabingas, Laura Gerstner, Stefanie Rachis

## Abstract

**Introduction:** Physician assistant (PA) programs face persistent challenges in recruiting and retaining clinical preceptors due to time constraints, administrative burden, lack of compensation, and limited training. Additional pressures, such as health care consolidation, program expansion, clinician burnout, and financial implications of paid clinical sites, further strain preceptorship capacity. This study examines motivators and barriers influencing clinicians’ willingness to precept PA students.

**Methods:** This mixed-methods study used snowball sampling to recruit current, former, and non-precepting PAs across North Carolina. Participants completed surveys with Likert-scale and open-ended items adapted from the 2011 National Survey of Physician Assistants. Four virtual focus groups, selected from survey respondents, underwent semi-structured interviews informed by Self-Determination Theory (SDT). Quantitative data were analyzed using descriptive statistics and ordinal logistic regression; qualitative data underwent thematic analysis with deductive SDT coding and inductive refinement. Triangulation integrated findings.

**Results:** Respondents (N = 158) represented diverse clinical experience. Top motivators included student quality (66%), program support (53%), and financial compensation (51%). Key barriers were student quality (61.29%), burnout (53.23%), and lack of compensation (46.77%). From the focused group discussion, four themes emerged: Student Quality, Financial Compensation, Non-Financial Incentives, and Administrative Support. Student preparedness acted as both motivator and barrier; compensation concerns focused on fairness.

**Discussion:** Preceptorship relies on relational and professional factors, student quality, recognition, and institutional alignment, rather than financial incentives alone. System inefficiencies, inadequate preparation, and misaligned compensation hinder engagement. Improving student readiness, enhancing institutional support, and implementing transparent, layered incentives may strengthen recruitment and retention.

## Introduction

Clinical preceptors play a vital role in physician assistant (PA) programs, serving as guides who create the nexus between classroom learning and clinical practice. Their guidance is invaluable in shaping the clinical and professional development of future PAs, imparting not only medical knowledge and skills but also essential aspects of patient care and ethical practice.

Despite their crucial role, the recruitment and retention of preceptors continue to pose significant obstacles for PA programs. As a result of the preceptor shortage, evaluation of the barriers to recruitment and retention have been considered. The lack of development and training was identified as a barrier to recruitment and retention of preceptors.^3,5,6,8^ Other barriers such as time constraints, financial compensation concerns, and administrative burdens often deter clinicians from taking on precepting roles.^1,5,6,8^

External factors which influence preceptor recruitment and retention include hospital mergers which alter affiliation agreements between hospitals and educational institutions as well as updated changes in the regulatory environment. This is further compounded by the expansion of existing accredited programs, as well as the increasing number of newly established health professional programs. There is increased faculty attrition in PA academia which include clinical coordinators leaving their positions, in part due to the increased stressors of this clinical site crisis.^2^ In addition, a soaring number of citations have been given to PA programs involving failing to meet Accreditation Review Commission on Education for the Physician Assistant’s (ARC-PA’s) standards pertaining to clinical education and not having a sufficient number of clinical sites.^4^

The impact of the COVID-19 pandemic on clinicians departing health care as well as re-evaluating work/life balance cannot be ignored. Overburdened health care systems and demands for increased productivity constrain potential preceptors from serving in a clinical preceptor role. A desire for financial incentives among some preceptors and health systems has added additional stress and may not be feasible for some institutions, especially in the long term. A growing proportion of PA programs are now paying for some or all clinical training sites, a trend that will likely worsen student debt and subsequently affect health care workforce capacity.^2,7^

Student debt will increase as preceptor payments are passed down to students with an increase in tuition costs at their individual programs.^7^

Inversely, there are positive motivating factors, such as potential professional fulfillment, the opportunity to give back to the profession, and the chance to pass the torch of knowledge to future health care providers to serve as compelling incentives for preceptors. Understanding these dynamics is essential for enhancing preceptor engagement and ensuring the continued success of PA education programs.

The purpose of this study is to examine the factors influencing clinically practicing PAs’ willingness to serve as preceptors for PA students in North Carolina and to determine whether these factors differ from those associated with unwillingness to precept. This study aims to identify both altruism-related and contextual motivators and barriers that inform preceptors’ decisions to engage in PA student mentorship. Additionally, this study seeks to describe the perceptions and experiences of PAs related to clinical preceptorship in PA education.

## Methods

This was a prospective, observational, mixed-methods study consisting of an initial survey, followed by focus group discussions with selected survey respondents. The study utilized a purposeful sampling approach to recruit Physician Assistant clinical preceptors across the state of North Carolina. A clinical preceptor was defined as “a PA who is primarily responsible for the clinical teaching of a physician assistant student for at least 4 weeks per year.” For the purposes of this study, this definition was expanded to also include PAs who have previously served as preceptors as well as those who are actively considering precepting in the future, regardless of whether they have yet hosted a student. This broader framing captured the full spectrum of clinicians engaged in, or contemplating engagement in, clinical education. Exclusion criteria included current PA students or other health professions students, PAs who practiced outside of the state of North Carolina, any PA who had never served as a PA preceptor and were not currently considering serving as a preceptor for PA students, or were unable to provide informed consent or complete the study procedures (e.g., due to language barriers or incomplete participation).

To further enhance the sample size and reach, a snowball sampling technique was implemented, whereby initial participants referred other qualified preceptors within their professional networks. Existing relationships with multiple North Carolina PA program clinical directors were leveraged to connect with the target population.

This study was reviewed and approved by the Advocate Health Wake Forest University School of Medicine Institutional Review Board, under protocol number IRB00122053. All participants in the survey and focus groups provided informed consent prior to data collection. Completed informed consents have been stored electronically within Qualtrics software (Qualtrics software, current version during the period of data collection, 2025) requiring secure login information.

### Survey

The survey consisted of a combination of Likert scales and open-ended questions. It was developed in Microsoft Forms, part of Microsoft Office 365, and input into Qualtrics for final dissemination via alumni listservs and professional networks. Likert scales were used to quantify the extent to which each factor influenced the decision to accept or decline preceptor roles (e.g., 5 = strongly agree to 1 = strongly disagree). Open-ended questions allowed respondents to elaborate on their responses and identify additional influencing factors.

The survey instrument was adapted from the Preceptor Experiences Survey Tool used in the 2011 National Survey of Physician Assistants.^14^ This tool was originally developed from March to August 2010 based on literature review and expert input from the PAEA Clinical Education Committee. Response options were drawn from previously published and unpublished instruments provided by professional PA organizations (AAPA, PAEA workshops).

The developed survey instrument was distributed online using emails attached to the principal investigators home institutional alumni listservs. The instrument utilized skip patterns to tailor questions based on respondents’ preceptor status. Open-ended questions were adapted from the four major themes identified during the literature review of prior qualitative research on perspectives of precepting students.^14^

### Focus Group Discussions

For focus group development, a structured guide was created to align with the research objectives and Self-Determination Theory (SDT) domains. Participants were recruited from survey completion—including current, former, and non-precepting clinicians. The focus group guide was piloted with a small, diverse group to assess question clarity and discussion flow. Participants readily understood and engaged with the questions, generating rich, relevant responses. Feedback confirmed the logical progression supported a natural conversational rhythm, and no substantive revisions were required.

Four one-hour virtual focus groups via Microsoft Teams, with 4-6 participants per group, were conducted to ensure representation across multiple clinical settings. These discussions allowed participants to explore and elaborate on their experiences and opinions in greater depth.

The mixed methods data analysis plan for this study integrated structured survey responses with qualitative insights to explore physician assistant precepting experiences. Initial quantitative analysis was conducted on survey data that included demographic variables (e.g., gender, years in practice, specialty), practice characteristics (e.g., setting, type of care), and Likert-scale ratings assessing incentives, barriers, workload, stress, and perceived support.

Descriptive statistics such as frequencies and percentages were used to summarize respondent characteristics, while ordinal logistic regression modeled predictors of willingness to precept, including compensation and institutional support. These statistical methods were selected due to the ordinal nature and non-normal distribution of Likert-scale data, ensuring robust and appropriate analysis.

Qualitative data was collected through open-ended responses and virtual focus groups. Thematic analysis was conducted on four transcripts derived from recorded Microsoft Teams, part of Microsoft 365, focus groups, employing a double-coding approach to enhance reliability and interpretive depth. Each transcript was independently reviewed by double coders who first familiarized themselves with the data, then generated initial codes based on recurring patterns, concepts, and participant language. Coders used a combination of inductive and deductive strategies, allowing themes to emerge organically while remaining attached to the study’s guiding questions. Thematic saturation was reached when additional transcripts no longer produced new codes, concepts, or insights. After double-coding the third and fourth transcripts, coders noted that all new data reinforced previously identified themes, with no novel patterns emerging. Saturation was confirmed during coder consensus meetings, where the team agreed that the themes were sufficiently rich, coherent, and theoretically grounded such that further data collection was unlikely to change the thematic structure. Qualitative analysis employed a deductive thematic approach grounded in Self-Determination Theory focusing on autonomy, competence, and relatedness as core motivational constructs.

Triangulation was used to connect quantitative and qualitative insights by comparing survey patterns, for example, the high value participants placed on compensation, with qualitative narratives that elaborated on those perceptions. In addition, a developmental design approach was used in which survey findings informed the construction of the focus group protocol. Specifically, the quantitative results guided the focus group questions, ensuring that the discussions probed more deeply into emerging areas of interest such as compensation fairness, workload, and motivations for teaching. This approach shaped the content of the discussion but did not influence the selection or makeup of focus group participants.

Insights from both the survey and focus groups were integrated to provide a comprehensive understanding of the factors influencing PA clinicians’ willingness to precept. The qualitative data served to validate and enrich the quantitative findings. Both the survey and focus group instruments underwent rigorous development and pilot testing to ensure validity and reliability.

## Results

A total of 158 participants completed the survey after being recruited using a snowball sampling approach. Because recruitment relied on participant referrals rather than a predefined invitation list, a conventional response rate could not be calculated. Survey participants represented a broad range of professional experience. *Table 1* illustrates the distribution of participants by years in practice, providing context for the range of clinical experience represented in the sample. This distribution reflects a balanced mix of early-, mid-, and late-career professionals, offering diverse perspectives across the continuum of clinical experience.

**Table 1:** Distribution of Survey Participants by Years in Practice.

| **Years in Clinical Practice** | **Respondents (n=158)** |
| --- | --- |
| 2 years or fewer | 17% |
| 3-5 years | 20% |
| 6-10 years | 17% |
| 11-15 years | 17% |
| More than 15 years | 29% |

### Perceived Importance of Precepting Incentives

Responses were grouped into two categories: *Important* (combining “Moderately Important” and “Extremely Important”) and *Not Important* (combining “Not at All Important” and “Slightly Important”). The top three incentives identified as most important were the **quality of students (66%), support from the PA program (53%)**, and **financial compensation (51%)**. *See Figure 2*. These factors emerged as the strongest motivators for PAs to engage in precepting.

**Figure 2:**
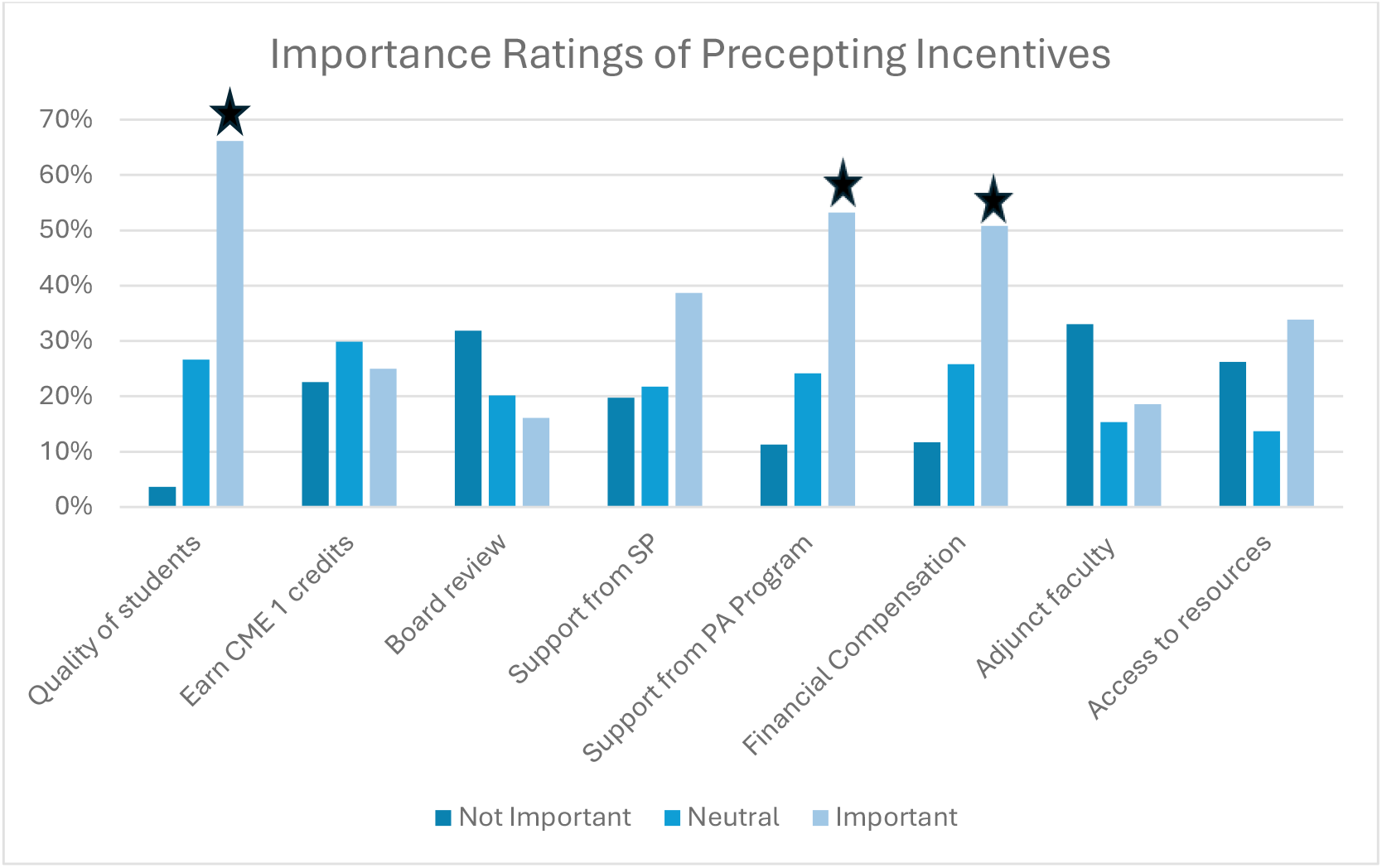
Importance Ratings of Precepting Incentives from Survey Instrument. Top three ratings designated with star symbol.

### Perceived Barriers to Precepting

Barriers to precepting were grouped into two categories: *Important* (combining “Moderately Important” and “Extremely Important”) and *Not Important* (combining “Not at All Important” and “Slightly Important”). The most frequently cited barriers were **quality of students (61.29%), burnout (53.23%)**, and **lack of financial compensation (46.77%)**, *See Figure 3*.

**Figure 3:**
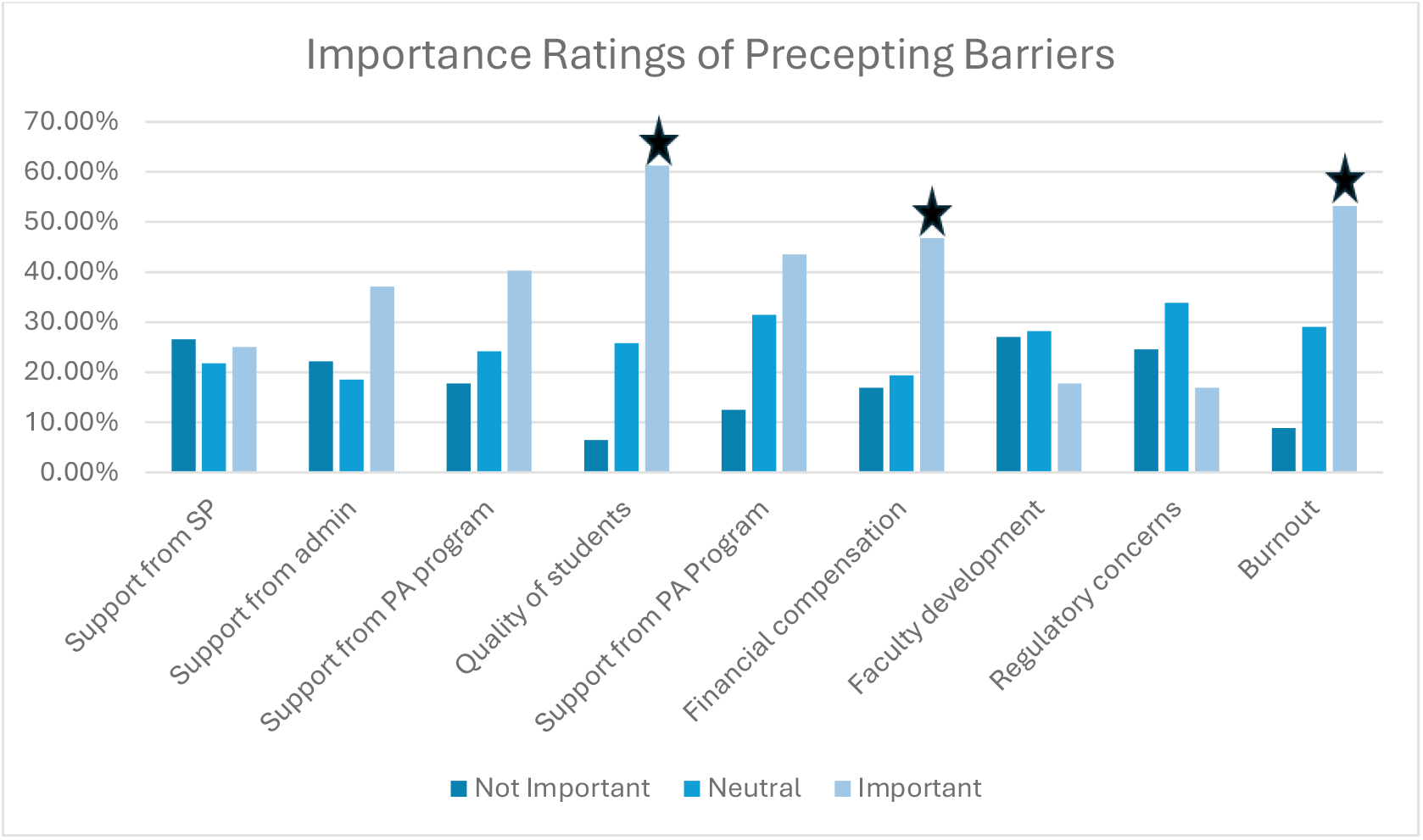
Importance Ratings of Precepting Barriers from Survey Instrument. Top three ratings designated with star symbol.

### Major Themes from Focus Groups

Four thematic findings emerged across focus group data. Themes were consistent across groups, with subtle variation by practice setting. Themes identified were Financial Compensation, Non-Financial Incentives, Administrative Support, and Student Quality. Summary of Themes is presented in *Table 2*.

**Table 2:** Summary of Qualitative Themes from Focus Group Discussions.

| Theme | Description & Evidence | Illustrative Evidence |
| --- | --- | --- |
| **Quality of Students** | Preceptors value initiative, professionalism, preparedness, and engagement. Students with a growth mindset are seen as assets. | “Initiative is really big...students who are self-motivated, who show that they want to be there...”<br>“If you have a bad crop of students, you really don't even have time to teach them...” |
| **Financial Compensation** | Compensation is appreciated but not the primary motivator. Concerns exist around fairness and distribution. | “Compensation is great, but I feel like I just enjoy precepting...for me at least, fine.” |
| **Non-Financial Incentives** | CME credits, adjunct status, access to university resources, and tax benefits are valued forms of recognition. | “Adjunct clinical status at the university...would give you access to the medical library, IRB, research facilities...” |
| **Institutional Support** | Support varies widely. Preceptors desire better scheduling, onboarding, and administrative assistance. | “If you could suggest one change to improve the preceptor process...giving someone time in their day is the best thing...” |

#### 1. Quality of Students

Across all groups, preceptors consistently cited the quality of students, including preparedness, initiative, professionalism, and engagement, as the most important factor influencing their willingness to precept.

#### 2. Financial Compensation

Compensation is appreciated but not the primary motivator. Many preceptors would continue without pay, but most agree that compensation is necessary in today’s environment. There is frustration when compensation is absorbed by institutions or supervising physicians rather than reaching the preceptor directly. Some organizations offer PTO or split payments, but lack of transparency and fairness is a recurring concern.

#### 3. Non-Financial Incentives

Category 1 CME credits for precepting are valued and sometimes serve as a substitute for financial compensation. Many preceptors expressed interest in adjunct or clinical faculty titles, which can support career advancement and provide access to institutional resources. Library access, UpToDate subscriptions, IRB involvement, and networking opportunities are seen as meaningful incentives. States like Maryland and Georgia offer tax credits for preceptors, which participants suggested as a model for North Carolina.

#### 4. Administrative and Institutional Support

Some preceptors feel well-supported by their institutions, while others report little to no recognition or assistance. Administrative turnover and poor communication can disrupt the precepting process. Preceptors desire more flexibility in scheduling and reduced patient loads when teaching, but most institutions do not accommodate this need. Smaller practices often lack clear processes for integrating students, leading to confusion and inefficiency.

### Integrated Findings: Barriers and Motivators to Precepting

Quantitative survey data and qualitative focus group themes revealed both convergence and divergence in perceived barriers to precepting. Convergent themes included time constraints, productivity pressures, and concerns about student quality. Notably, the quality of students emerged as a dual construct: while high-quality learners were cited as a key incentive, concerns about inadequate preparation surfaced as a recurring barrier.

Divergence was observed in the emotional tone and depth of responses. While survey participants reported moderate satisfaction with precepting experiences, focus group discussions uncovered deeper accounts of emotional labor, role strain, and burnout.

From these integrated findings, two meta-inferences emerged. First, recognition and relational support, such as feeling valued by programs and connected to learners, carry greater motivational weight than financial incentives alone. Second, sustained engagement in precepting appears to depend on institutional alignment and student preparation, reinforcing the need for coordinated support across administrative, academic, and clinical domains.

A concept that did not meet thematic threshold across qualitative data was the concept of *precepting as long-term influence*. Several participants described teaching not only as a professional responsibility but as a strategic avenue to influence future practice norms and address systemic inequities. For these preceptors, mentorship served as a mechanism to instill values of equity, professionalism, and patient-centered care in the next generation, positioning clinical education as a form of long-term change-making within the health care system.

## Discussion

Among all factors examined, student quality emerged as the primary determinant of preceptors’ willingness to precept. More than financial incentives or institutional support, the preparedness, professionalism, and initiative of learners emerged as the decisive factor influencing preceptorship engagement. While survey data confirmed well-documented barriers, such as time constraints, lack of compensation, and productivity pressures, qualitative insights illustrated the emotional labor and misalignment between preceptors’ values and institutional structures. Together, these findings emphasize the need to reconceptualize precepting as a relational and identity-affirming endeavor, rather than a transactional task. The data from this study supports the hypothesis that more than financial factors play a central role in shaping preceptors’ willingness to mentor PA students.

Preceptors consistently described their motivation as rooted in professional identity, mentorship, and the desire to shape future clinicians. This aligns with Self-Determination Theory (SDT), which posits that autonomy, competence, and relatedness are foundational to sustained engagement.^9^ However, institutional demands, particularly productivity metrics and lack of protected time, constrained autonomy. Varying levels of student readiness complicated judgments of competence, while insufficient acknowledgment weakened relational connections. These tensions suggest that precepting disengagement may stem less from workload volume and more from the wearing down of motivational drivers central to SDT.

These findings mirror prior research in medical and nursing education that identifies time constraints, lack of compensation, and insufficient institutional support as persistent barriers to precepting(^10−12^) This study expands the current literature by centering the emotional and identity dimensions of precepting, areas often neglected in survey-only approaches. The integration of qualitative data reveals how systemic pressures interact with personal values, offering a richer understanding of preceptor experience.

The dual role of student quality, as both a motivator when high and a barrier when lacking, deepens the understanding of prior findings. Previous studies have noted that inadequate student preparation can exacerbate preceptor workload and contribute to burnout.(^10−12^) The present findings reinforce this tension, suggesting that student readiness is not merely a background factor but a pivotal determinant of whether precepting is experienced as rewarding or burdensome.

The mixed views on financial compensation also echo prior literature. Notably, focus group participants emphasized that financial incentives were not reaching preceptors directly. Instead, compensation, when offered, was often routed to supervising physicians, departments, or organizations, leaving individual preceptors without tangible acknowledgment of their labor. This disconnect reinforces the perception of precepting as undervalued and contributes to feelings of distributive injustice. The lack of direct financial recognition compounds emotional labor and may exacerbate burnout, particularly when preceptors are expected to absorb additional responsibilities without institutional reciprocity. Some states have shown that stipends or tax credits can increase recruitment of preceptors as compensation alone has rarely been sufficient to ensure long-term engagement.(^2,13^) The participants’ emphasis on fairness, transparency, and direct benefit to the preceptor highlights systemic issues in how compensation is structured.

Non-financial incentives such as Category I CME credits, adjunct titles, and access to academic resources were consistently valued. Tiered incentive systems, where benefits scale with the number of students precepted, could encourage broader participation. Creative incentives, such as career development opportunities, institutional perks, or recognition for recruiting future employees, were also proposed. These forms of recognition not only support professional development but also strengthen the relational ties between preceptors and academic institutions. The implication is that programs seeking to expand preceptorship capacity should consider layered incentive models that combine modest financial support with meaningful professional recognition.

Institutional support emerged as another critical dimension. Consistent with prior literature, participants described variability in administrative processes, communication, and workload adjustments.(^10−12^) The lack of structural flexibility, particularly in smaller practices, suggests that institutional alignment remains a weak link in the precepting environment. Without coordinated support, even highly motivated preceptors may disengage due to competing productivity pressures.

These findings highlight that preceptorship is sustained less by transactional incentives and more by relational, professional, and institutional alignment. PA programs that prioritize student preparation, provide visible recognition, and foster transparent compensation structures are more likely to cultivate enduring preceptor engagement. In addition, the emergent theme of precepting as long-term influence suggests that many clinicians view teaching as a means of shaping future practice norms and advancing equity. This positions preceptorship not only as a workforce necessity but also as a mechanism for long-term cultural change in health care.

While the mixed-methods design strengthens the validity of findings, several limitations warrant consideration. The study was conducted in a single state, which may limit generalizability to regions with different policy environments or institutional structures. Survey responses may also reflect self-selection bias, with more engaged or motivated preceptors choosing to participate, potentially skewing results towards those with stronger opinions about precepting. While survey participants reported moderate satisfaction with precepting overall, focus group discussions revealed deeper accounts of emotional labor, role strain, and burnout. These qualitative insights suggest that the affective dimensions of precepting may be underrepresented in structured survey instruments. This highlights the importance of integrating qualitative approaches to better understand the lived experiences of preceptors. Finally, social desirability bias may have influenced focus group responses, with participants potentially moderating negative feedback in group settings.

Building on these findings, future research could evaluate the effectiveness of recognition and support interventions on preceptor retention. Institutional readiness for structural change, particularly around protected time and workload redistribution, warrants further exploration as well. Additionally, examining student-preceptor dynamics and their influence on teaching satisfaction could inform onboarding and matching processes. Developing metrics for relational and emotional dimensions of precepting would enable programs to assess and respond to preceptor needs more holistically. Finally, a deeper understanding of what constitutes well versus poorly prepared students, including evolving professionalism norms across generations, may explain how perceptions differ among preceptors at varying career stages. Such insights could inform both student preparation strategies and faculty development, while also offering valuable guidance to accrediting bodies such as PAEA and ARC-PA in shaping standards that align with present workforce expectations.

## Conclusion

This study emphasizes that the sustainability of PA preceptorship centers on more than financial incentives. Student quality, recognition, and institutional support emerged as the most powerful drivers of engagement, while inadequate preparation and systemic inefficiencies remain persistent barriers. For policymakers and program leaders, the takeaway is clear: investment in student readiness, transparent incentive structures, and institutional alignment will yield greater returns than financial compensation alone. By reframing preceptorship as both a professional responsibility and a form of shaping the future workforce, programs can foster a culture where teaching is valued not only as service but as a strategic contribution to the future of health care.

## Data Availability

All data produced in the present study are available upon reasonable request to the authors

## Acknowledgements

This manuscript was created as part of graduation requirements for the DMSc program of Wake Forest University School of Medicine Physician Assistant Studies.

A pre-press copy of this manuscript has been posted to the medRxiv.org pre-print server.

